# Sex Differences in PTSD Risk Among Autistic Individuals: A Population-Based Matched Cohort Study

**DOI:** 10.64898/2026.03.31.26349863

**Authors:** Shelby Smout, Seulgi Jung, Veerle Bergink, Behrang Mahjani

## Abstract

**Objective:** Autistic individuals may face elevated risk for PTSD, yet the degree to which this risk differs by sex remains unknown. We examined the association between autism and incident PTSD, characterized sex differences in risk, identified high-risk subgroups, and described post-diagnosis clinical trajectories.

**Method:** We conducted a population-based matched cohort study using Swedish national registers. Individuals born 1990 through 2010 were followed from age 6 years through December 31, 2017. Autistic individuals (N=42,862) were matched 1:10 to controls (N=412,251) on sex and birth year. Cox proportional hazards regression estimated hazard ratios (HRs) for incident PTSD. Among those who developed PTSD, we compared care utilization, hospitalization rates, and persistence of care contacts.

**Results:** During mean follow-up of 5.1 years, 401 autistic individuals (0.9%) and 903 controls (0.2%) developed PTSD (incidence rates: 18.3 vs 4.2 per 10,000 person-years). Autism was associated with 4.4-fold increased PTSD risk (HR=4.37; 95% CI, 3.93-4.86). Risk was higher among females (HR=4.79) than males (HR=3.39; P interaction=.006). Among autistic individuals, comorbid ADHD conferred additional risk (HR=1.38; 95% CI, 1.14-1.68). Ten-year cumulative incidence reached 6.0% among autistic females with ADHD. Autistic individuals with PTSD had higher care utilization (mean visits: 5.0 vs 3.9; P<.001), more psychiatric hospitalizations (27.9% vs 19.8%; P=.002), and more persistent courses (24.8% vs 12.3% with contacts in all 3 post-diagnosis years; P=.001).

**Conclusion:** Autism is associated with substantially elevated PTSD risk, particularly among females with comorbid ADHD. When PTSD occurs, autistic individuals experience more severe and persistent clinical courses, supporting targeted screening and sustained follow-up.

## INTRODUCTION

Posttraumatic stress disorder (PTSD) is a debilitating psychiatric condition that develops following exposure to traumatic events and is characterized by intrusive memories, avoidance behaviors, negative alterations in cognition and mood, and hyperarousal.^1^ While PTSD affects approximately 3.9% of the general population worldwide,^2^ females are disproportionately affected, developing PTSD at approximately twice the rate of men, even after accounting for differences in trauma exposure.^2,3^ Emerging evidence suggests that individuals with autism spectrum disorder (ASD) also face substantially elevated risk.^4–7^ A nationwide cohort study from Taiwan examining over 15,000 children and adolescents with ASD found significantly elevated risk for both acute stress disorder and PTSD compared to matched controls.^8^ Similar elevations in PTSD risk have been documented among individuals with attention-deficit/hyperactivity disorder (ADHD),^9,10^ which co-occurs in approximately 40–70% of autistic individuals. This co-occurrence is important to note as it may compound PTSD vulnerability through multiple neurobiological and psychosocial pathways, and its interaction with sex warrants particular consideration.^11^

Several mechanisms may contribute to elevated PTSD vulnerability among autistic individuals, operating through both increased trauma exposure and heightened vulnerability to traumatization. Autistic individuals experience substantially higher rates of adverse life events, including bullying, social exclusion, and various forms of victimization.^12,13^ Deficits in social cognition, such as difficulty identifying inappropriate behavior, detecting violations of social norms, and recognizing one’s own discomfort, may increase susceptibility to interpersonal trauma by impairing the recognition of dangerous situations.^14^ In addition to elevated trauma exposure, neurodevelopmental characteristics may lower the threshold for PTSD development, meaning events not typically traumatic for neurotypical individuals may be sufficient to trigger PTSD in autistic individuals. Altered sensory reactivity, particularly pronounced in autism, may intensify the subjective experience of potentially traumatic events, with heightened sensory responsivity predicting PTSD symptomatology.^15^ Difficulties with emotion regulation, documented in both autism and ADHD, may impede adaptive coping strategies following trauma exposure.^16^ Chronic stress from lifelong challenges in social interactions and executive functioning may further deplete psychological reserves, as chronic stressor exposure itself represents an independent PTSD risk factor.^17^ Whether these mechanisms operate differently across sexes, and to what degree they explain observed sex differences in PTSD risk among autistic individuals, remains unknown.

Despite the well-established female predominance of PTSD in the general population, a striking and clinically important pattern emerges in autism. While autism is more commonly diagnosed in males, emerging evidence suggests that PTSD among autistic individuals may show a female predominance, indicating that females bear a disproportionate burden despite being diagnosed with autism less frequently.^18^ The mechanisms underlying this sex reversal remain poorly understood. Multiple factors have been proposed to contribute to elevated risk, specifically among autistic females. First, autistic females experience diagnostic delays averaging 2 to10 years compared to males, potentially prolonging exposure to adverse experiences during vulnerable developmental periods without appropriate supports.^19–21^ Second, females may present with a distinct autism phenotype characterized by better social camouflaging, less obvious repetitive behaviors, and more internalizing symptoms, complicating both trauma recognition and PTSD diagnosis.^22,23^ Third, the frequent co-occurrence of autism and ADHD introduces additional complexity, as diagnostic delays are further exacerbated when both conditions are present,^24^ and ADHD itself independently predicts PTSD risk.^11^ Yet these explanations remain largely untested in population-based samples. Furthermore, prior studies have relied primarily on cross-sectional designs, clinical samples, or self-report measures, precluding estimation of sex-stratified incidence rates and limiting generalizability.^4,5^ Whether female sex and ADHD comorbidity exert independent effects on PTSD risk or interact remains unknown, yet understanding their combined influence is essential for developing targeted screening and intervention approaches.

The present study addressed these gaps using Swedish national registers, which provide comprehensive, longitudinal psychiatric diagnostic data for the entire population. We examined three primary questions: (1) What is the association between autism and incident PTSD, and does this association differ by sex? (2) Do female sex and ADHD comorbidity exert independent or interactive effects on PTSD risk among autistic individuals? (3) Among those who develop PTSD, do autistic individuals experience different clinical courses than non-autistic individuals? We hypothesized that autism would be associated with elevated PTSD risk, that this association would be stronger among females, that female sex and ADHD would independently predict risk within the autism group, and that autistic individuals with PTSD would demonstrate more severe and persistent clinical courses.

## METHOD

### Study Design and Population

We conducted a nationwide matched cohort study using Swedish national registers. The study population comprised all individuals born in Sweden between January 1, 1990, and December 31, 2010, identified through the Swedish Medical Birth Register, which captures more than 98% of all births.^25^ Participants entered the cohort at age 6 years to allow adequate time for autism diagnosis while capturing the full range of PTSD onset ages. We excluded individuals who emigrated or died before age 6 years. Follow-up continued through December 31, 2017, death, or emigration, whichever occurred first. The Regional Ethical Review Board in Stockholm approved this study (Dnr 2013/862-31/5). Swedish law permits register-based research without individual consent when data are anonymized.

### Data Sources

Psychiatric diagnoses were ascertained from the National Patient Register, which includes all psychiatric inpatient admissions since 1973 and outpatient specialist visits since 2001.^26^ To ensure consistent diagnostic coding, we restricted analyses to diagnoses recorded from January 1, 1997, onward, when ICD-10 implementation was complete. The register captures diagnoses in both primary and up to 30 secondary positions, and we included diagnoses from any position. Family relationships were established through the Multi-Generation Register, which links individuals to their biological parents.^27^ Medication dispensations were obtained from the Swedish Prescribed Drug Register, which has complete national coverage since July 2005.^28^ Dates of death and emigration were ascertained from the Cause of Death Register and the Register of the Total Population, respectively.

### Exposure Definition

Autism spectrum disorder was defined as at least one recorded diagnosis with ICD-10 codes F84.0 (childhood autism), F84.1 (atypical autism), F84.5 (Asperger syndrome), F84.8 (other pervasive developmental disorders), or F84.9 (pervasive developmental disorder, unspecified). We excluded F84.2 (Rett syndrome) and F84.3 (childhood disintegrative disorder), which are etiologically distinct conditions.^29^ The index date for autistic individuals was the date of first autism diagnosis recorded at or after age 6 years. Swedish register-based autism diagnoses have demonstrated high validity, with positive predictive values exceeding 90%.^29^

### Outcome Definition

The primary outcome was incident PTSD, defined as the first recorded diagnosis with ICD-10 code F43.1 after the index date. Diagnoses were identified from any diagnostic position in the National Patient Register. Individuals with PTSD diagnosed before the index date were excluded from both exposure groups to ensure analysis of incident cases.

### Covariates

Covariates assessed at or before the index date included ADHD (ICD-10 F90.x), intellectual disability (ICD-10 F70-F79), and parental psychiatric history. Parental psychiatric history was defined as any F-chapter diagnosis recorded in the National Patient Register for biological mothers or fathers identified through the Multi-Generation Register. Sex and birth year were obtained from the Medical Birth Register.

### Matching

Each autistic individual diagnosed at or after age 6 years was matched to 10 controls without autism. Matching was performed on sex (exact) and birth year (nearest neighbor) using the MatchIt package in R.^30^ Controls were assigned the same index date as their matched case to ensure comparable follow-up periods. This matching ratio was selected to maximize statistical power while maintaining computational feasibility.

### Statistical Analysis

Follow-up began at the index date and continued until PTSD diagnosis, emigration, death, or December 31, 2017. Incidence rates were calculated as PTSD events per 10,000 person-years. Cumulative incidence was estimated using the Kaplan-Meier method (1 minus survival probability) at 5 and 10 years.

Cox proportional hazards regression estimated hazard ratios (HRs) and 95% confidence intervals (CIs) for the association between autism and incident PTSD. Age served as the underlying time scale, with participants entering at age at index and exiting at age at event or censoring. All models were stratified by matched set to account for the matched design. Robust standard errors were clustered on biological mother to account for sibling correlation.

We fit sequential models adjusting for ADHD, intellectual disability, and parental psychiatric history to quantify the degree to which each covariate attenuated the autism-PTSD association. Effect modification was assessed through stratified analyses and multiplicative interaction terms for sex, intellectual disability, and ADHD status. Within the autism group, we examined joint effects of sex and ADHD using a four-level categorical variable (male without ADHD as reference, male with ADHD, female without ADHD, female with ADHD).

Number needed to screen (NNS) was calculated as 100 divided by the 10-year absolute risk difference between each risk stratum and the reference group (autistic males without ADHD).

### Exploratory Analyses

In exploratory analyses restricted to autistic individuals, we examined whether age at autism diagnosis and autism subtype contributed to PTSD risk and whether they explained the observed sex difference. Age at autism diagnosis was modeled continuously (per year) and categorically (6-9 years as reference, 10-12, 13-15, and 16+ years). To quantify the degree to which diagnostic timing explained the sex difference in PTSD risk, we compared the HR for female sex before and after adjustment for age at diagnosis. Effect modification by sex was assessed through sex-stratified models and a multiplicative interaction term.

For autism subtype analyses, we classified individuals as having Asperger syndrome (ICD-10 F84.5) versus other autism spectrum diagnoses (F84.0, F84.1, F84.8, F84.9) based on first recorded diagnosis. Although Asperger syndrome has since been integrated into the broader autism spectrum disorder diagnosis in DSM-5, this classification remains in ICD-10 and historically characterized individuals without intellectual disability or early language delay, thus providing a proxy for examining whether PTSD risk varied by autism presentation or support needs. We fit models adjusted for sex and birth year only, followed by fully adjusted models including ADHD, intellectual disability, and parental psychiatric history. Sex-stratified analyses examined whether subtype effects differed by sex, with effect modification assessed via interaction term.

### Post-PTSD Clinical Course Analysis

Among individuals who developed PTSD, we compared clinical course between autistic individuals and controls. Care utilization was measured as the total number of PTSD-coded healthcare visits (ICD-10 F43.1 in any diagnostic position) recorded after the initial diagnosis. Maximum care-setting severity was classified hierarchically based on care type: outpatient specialist care only, general psychiatric inpatient care, or specialized psychiatric inpatient care, based on the highest level of care received during follow-up.

Persistence was operationalized as the proportion with any PTSD-coded contact in each of years 1 through 5 following diagnosis. Among individuals with at least 3 years of post-diagnosis follow-up, we calculated the proportion with contacts in all three years. Groups were compared using Wilcoxon rank-sum tests for continuous variables, chi-square tests for categorical variables, and Fisher exact tests when expected cell counts were below 5.

### Sensitivity Analyses

To examine whether ADHD medication might be associated with PTSD risk among autistic individuals with ADHD, we conducted a secondary analysis comparing those who received at least two ADHD medication dispensations (ATC codes N06BA01, N06BA02, N06BA04, N06BA07, N06BA09, N06BA11, N06BA12) within the first year after ADHD diagnosis versus those who did not. This fixed exposure window approach avoids immortal time bias that would arise from defining medication status over the entire follow-up period.

All analyses were conducted in R version 4.2 using the survival^31^, survminer^32^, data.table^33^, and MatchIt^30^ packages. Two-sided P values below .05 were considered statistically significant.

## RESULTS

### Cohort Characteristics

The birth cohort comprised 2,091,180 individuals, of whom 49,395 (2.4%) received an autism diagnosis. After applying inclusion criteria (autism diagnosis at or after age 6, no prevalent PTSD), 42,862 autistic individuals were matched 1:10 to 412,251 controls (Table 1). Matching achieved balance on sex (32.7% vs 32.5% female) and birth year (median 1998 in both groups). As expected, autistic individuals had substantially higher prevalence of ADHD (42.5% vs 2.6%), intellectual disability (9.6% vs 0.6%), and parental psychiatric history (44.9% vs 24.3%; all P<.001).

**Table 1.** Baseline Characteristics of Autistic Individuals and Matched Controls.

| Characteristic | Autism (N=42,862) | Controls (N=412,251) | P Value |
| --- | --- | --- | --- |
| Female, No. (%) | 14,015 (32.7) | 133,923 (32.5) | .38 |
| Birth year, median (IQR) | 1998 (1994-2002) | 1998 (1994-2002) | .19 |
| Age at index, mean (SD), y | 14.0 (4.7) | 13.9 (4.7) | <.001 |
| Follow-up, mean (SD), y | 5.1 (4.0) | 5.3 (3.9) | <.001 |
| ADHD, No. (%) | 18,222 (42.5) | 10,678 (2.6) | <.001 |
| Intellectual disability, No. (%) | 4,129 (9.6) | 2,347 (0.6) | <.001 |
| Parental psychiatric history, No. (%) | 19,235 (44.9) | 100,066 (24.3) | <.001 |
| PTSD events, No. (%) | 401 (0.9) | 903 (0.2) | <.001 |
| Incidence rate per 10,000 person-years | 18.3 | 4.2 |  |

Abbreviations: ADHD, attention-deficit/hyperactivity disorder; IQR, interquartile range; PTSD, posttraumatic stress disorder.

### Incidence of PTSD

During a mean follow-up of 5.1 years (range, 0-21.9), 401 autistic individuals (0.9%) and 903 controls (0.2%) received a PTSD diagnosis. Incidence rates were 18.3 per 10,000 person-years among autistic individuals and 4.2 per 10,000 person-years among controls (Figure S1). Five-year cumulative incidence was 0.9% (95% CI, 0.8%-1.0%) in the autism group and 0.2% (95% CI, 0.2%-0.2%) in controls. By 10 years, cumulative incidence reached 1.8% (95% CI, 1.6%-2.1%) among autistic individuals and 0.4% (95% CI, 0.4%-0.5%) among controls.

Among individuals who developed PTSD, median time from index date to diagnosis was shorter in autistic individuals than controls (3.4 years [IQR, 1.5-5.6] vs 4.0 years [IQR, 2.0-6.8]; P<.001).

### Association Between Autism and PTSD

In the primary analysis, autism was associated with a 4.4-fold increased risk of incident PTSD (HR=4.37; 95% CI, 3.93-4.86; Table 2). This association was partially attenuated after adjustment for ADHD (HR=3.32; 95% CI, 2.88-3.82) and parental psychiatric history (HR=3.67; 95% CI, 3.28-4.11), suggesting these factors partially mediate or confound the association. Adjustment for intellectual disability slightly strengthened the association (HR=4.64; 95% CI, 4.16-5.17), consistent with intellectual disability being associated with lower PTSD ascertainment.

**Table 2.** Hazard Ratios for PTSD Risk by Adjustment Model.

| Model | Autism HR (95% CI) | Covariate HR (95% CI) |
| --- | --- | --- |
| Unadjusted | 4.37 (3.93-4.86) | — |
| Adjusted for ADHD | 3.32 (2.88-3.82) | 2.05 (1.68-2.50) |
| Adjusted for intellectual disability | 4.64 (4.16-5.17) | 0.45 (0.29-0.70) |
| Adjusted for parental psychiatric history | 3.67 (3.28-4.11) | 2.43 (2.16-2.73) |

Abbreviations: ADHD, attention-deficit/hyperactivity disorder; HR, hazard ratio. All models stratified by matched set with robust standard errors clustered on biological mother.

### Effect Modification by Sex

The autism-PTSD association differed significantly by sex (P for interaction = .006; Table 3). Although autism was associated with elevated PTSD risk in both sexes, the association was stronger among females (HR=4.79; 95% CI, 4.24-5.41) than males (HR=3.39; 95% CI, 2.74-4.20). Absolute risk differences were also more pronounced among females: 10-year cumulative incidence was 4.8% (95% CI, 4.1%-5.5%) among autistic females compared with 1.0% (95% CI, 0.9%-1.1%) among female controls, whereas among males, cumulative incidence was 0.6% (95% CI, 0.4%-0.8%) in autistic individuals versus 0.2% (95% CI, 0.2%-0.2%) in controls.

**Table 3.** Stratified Analyses of PTSD Risk Among Autistic Individuals.

| Stratum | No. | Events | 10-y CI, % | HR (95% CI) | P |
| --- | --- | --- | --- | --- | --- |
| **By sex** |  |  |  |  | .006 |
| Female | 14,015 | 308 | 4.8 (4.1-5.5) | 4.79 (4.24-5.41) | <.001 |
| Male | 28,847 | 93 | 0.6 (0.4-0.8) | 3.39 (2.74-4.20) | <.001 |
| **By intellectual disability** |  |  |  |  | .20 |
| With ID | 4,129 | 19 | 0.5 | 1.88 (1.19-2.98) | .007 |
| Without ID | 38,733 | 382 | 1.0 | 4.68 (4.20-5.22) | <.001 |
| **By ADHD (within autism)** |  |  |  |  | .001 |
| With ADHD | 18,222 | 168 | 2.1 (1.6-2.5) | 1.38 (1.14-1.68) | .001 |
| Without ADHD (ref) | 24,640 | 233 | 1.7 (1.5-2.0) | 1 [Reference] |  |

Abbreviations: ADHD, attention-deficit/hyperactivity disorder; CI, cumulative incidence; HR, hazard ratio; ID, intellectual disability. P values in the stratum header rows are for multiplicative interaction terms.

Notably, among individuals who developed PTSD, 76.8% of autistic cases were female compared with 70.4% of control cases. This female predominance represents a striking reversal of the male-predominant sex ratio typically observed in autism (67.3% male in our cohort).

### Effect Modification by Intellectual Disability

Among autistic individuals, those with intellectual disability had lower PTSD incidence than those without (0.5% vs 1.0% at 10 years). The HR for autism versus controls was attenuated in the intellectual disability stratum (HR=1.88; 95% CI, 1.19-2.98) compared with those without intellectual disability (HR=4.68; 95% CI, 4.20-5.22), though the interaction did not reach statistical significance (P=.20). This pattern may reflect diagnostic overshadowing, communication barriers affecting PTSD diagnosis, or genuine differences in trauma exposure or processing.

### ADHD as a Risk Factor Within Autism

Among autistic individuals, comorbid ADHD was associated with 38% increased PTSD risk (HR=1.38; 95% CI, 1.14-1.68; P=.001). Ten-year cumulative incidence was 2.1% (95% CI, 1.6%-2.5%) among autistic individuals with ADHD and 1.7% (95% CI, 1.5%-2.0%) among those without ADHD. ADHD was highly prevalent in the autism group (42.5%), meaning this risk factor affected a substantial proportion of the population.

### Joint Effects of Sex and ADHD

Given the independent effects of female sex and ADHD on PTSD risk within autism, we examined their joint effects (Table 4; Figure S2). Risk stratification revealed marked heterogeneity. With autistic males without ADHD as reference, HRs were 1.85 (95% CI, 1.23-2.77) for males with ADHD, 7.74 (95% CI, 5.60-10.69) for females without ADHD, and 10.69 (95% CI, 7.61-15.02) for females with ADHD.

**Table 4.** Clinical Risk Stratification by Sex and ADHD Status Among Autistic Individuals.

| Group | No. | Events | IR | 10-y CI, % | HR (95% CI) | NNS |
| --- | --- | --- | --- | --- | --- | --- |
| Male without ADHD | 16,007 | 46 | 4.8 | 0.5 (0.3-0.7) | 1 [Reference] | — |
| Male with ADHD | 12,840 | 47 | 8.1 | 0.7 (0.5-0.9) | 1.85 (1.23-2.77) | 670 |
| Female without ADHD | 8,633 | 187 | 42.5 | 4.3 (3.6-5.1) | 7.74 (5.60-10.69) | 26 |
| Female with ADHD | 5,382 | 121 | 56.1 | 6.0 (4.5-7.4) | 10.69 (7.61-15.02) | 18 |

Abbreviations: ADHD, attention-deficit/hyperactivity disorder; CI, cumulative incidence; HR, hazard ratio; IR, incidence rate per 10,000 person-years; NNS, number needed to screen over 10 years (calculated as 100/risk difference vs reference group).

Ten-year cumulative incidence was 6.0% (95% CI, 4.5%-7.4%) among autistic females with ADHD, representing approximately 1 in 17 individuals. The incidence rate in this group (56.1 per 10,000 person-years) was more than 11-fold higher than among autistic males without ADHD (4.8 per 10,000 person-years). The NNS to identify one additional PTSD case among autistic females with ADHD (compared with males without ADHD) was 18.

### Exploratory Analyses: Age at Diagnosis and Autism Subtype

Age at autism diagnosis was associated with PTSD risk among autistic individuals. Each additional year of age at diagnosis conferred 9% increased risk (HR=1.09; 95% CI, 1.06-1.13; P<.001), with categorical analyses revealing a dose-response relationship: compared with diagnosis at ages 6-9 years, diagnosis at 13-15 years was associated with HR=1.93 (95% CI, 1.21-3.09) and diagnosis at 16+ years with HR=2.35 (95% CI, 1.48-3.72) (Supplemental Table 1A). Females were diagnosed significantly later than males (mean 15.0 vs 13.5 years; P<.001), with 73.2% of females diagnosed after age 12 compared with 57.7% of males. However, this diagnostic delay explained very little of the sex difference in PTSD risk. Adjusting for age at diagnosis attenuated the female HR from 6.63 (95% CI, 5.26-8.37) to 6.15 (95% CI, 4.87-7.77), representing only 4% attenuation. The effect of diagnostic age on PTSD risk was equivalent in both sexes (HR=1.09 for both; P for interaction=.87).

Individuals with Asperger syndrome (F84.5) had higher crude PTSD incidence than those with other autism diagnoses (24.5 vs 14.0 per 10,000 person-years). Asperger syndrome was associated with elevated PTSD risk when adjusted for sex and birth year only (HR=1.33; 95% CI, 1.09-1.62; P=.005), but this association was attenuated and no longer statistically significant after full covariate adjustment (HR=1.16; 95% CI, 0.95-1.42; P=.15). In sex-stratified analyses, the Asperger-associated elevation was observed among females (HR=1.38; 95% CI, 1.10-1.73; P=.005) but not males (HR=1.17; 95% CI, 0.77-1.76; P=.47), though the interaction was not significant (P=.45) (Supplemental Table 1B).

### Post-PTSD Clinical Course

Among individuals who developed PTSD (N=401 autistic, N=903 controls), autistic individuals demonstrated more intensive and persistent clinical courses (Table 5). Mean number of PTSD-coded visits after diagnosis was 5.0 (SD=6.1) among autistic individuals compared with 3.9 (SD=5.7) among controls (P<.001).

**Table 5.**
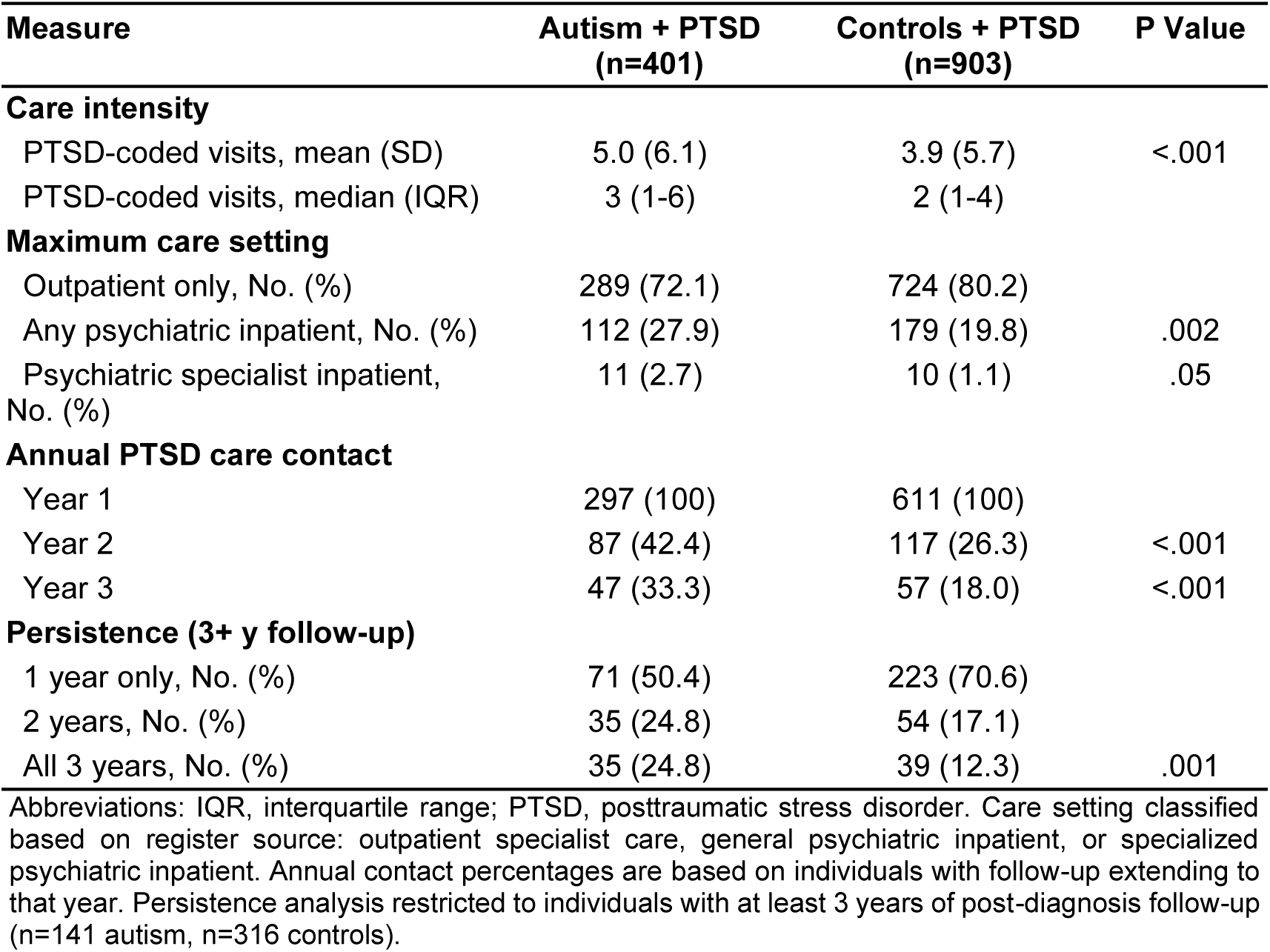
PTSD Clinical Course After Diagnosis.

Maximum care-setting severity also differed between groups. Among autistic individuals with PTSD, 27.9% received inpatient psychiatric care at some point during follow-up, compared with 19.8% of controls (P=.002). Compulsory psychiatric admission under the Swedish Mental Health Act occurred in 2.7% of autistic individuals and 1.1% of controls (P=.05).

Persistence of PTSD-related care contacts was markedly higher among autistic individuals. In year 2 after diagnosis, 42.4% of autistic individuals had PTSD-coded contacts compared with 26.3% of controls (P<.001). This differential persisted through year 3 (33.3% vs 18.0%; P<.001). Among those with at least 3 years of follow-up after PTSD diagnosis (N=141 autistic, N=316 controls), 24.8% of autistic individuals had PTSD-coded contacts in all three years compared with 12.3% of controls (P=.001).

## DISCUSSION

In this population-based matched cohort study, we found that autism was associated with a 4.4-fold increased risk of incident PTSD. Most strikingly, this risk was markedly higher among females than males, with autistic females accounting for 77% of PTSD cases despite comprising only 33% of the autistic cohort. The association was partially explained by ADHD comorbidity and parental psychiatric history. Among autistic individuals, female sex and ADHD were independently associated with elevated risk, with autistic females with ADHD experiencing more than 10-fold higher risk than autistic males without ADHD. Critically, when PTSD occurred, autistic individuals demonstrated more severe and persistent clinical courses, with higher rates of psychiatric hospitalization and sustained care needs over multiple years.

The sex difference in PTSD risk among autistic individuals merits particular attention. While autism is more commonly diagnosed in males (67% in our cohort), PTSD among autistic individuals was predominantly female (77% of cases). This sex reversal suggests that autistic females face unique vulnerabilities. Potential mechanisms include higher rates of sexual victimization among autistic females,^18,23^ camouflaging behaviors that may increase psychological strain,^23^ and possible differences in trauma processing related to sex-differentiated autism phenotypes.^22^ Current research estimates that two-thirds of autistic women who experience sexual victimization are first assaulted before age 18, highlighting the importance of early trauma exposure in understanding PTSD risk.^34^ Our exploratory analyses shed light on which commonly hypothesized factors do, and do not, account for this disparity. Consistent with prior research, females were diagnosed on average 1.5 years later than males, and later diagnosis was associated with increased PTSD risk (HR=1.09 per year). However, adjustment for diagnostic timing attenuated the female excess by only 4%, suggesting that delayed diagnosis does not uniquely harm females but rather that females simply experience more of it. Similarly, elevated crude PTSD incidence among those with Asperger syndrome was largely explained by confounding factors after full covariate adjustment. These null findings strengthen the inference that unmeasured factors, particularly differential trauma exposure, likely drive the sex disparity, and underscore the importance of PTSD screening among autistic females, who may be at especially high risk despite receiving less clinical attention than autistic males.

The magnitude of association observed here (HR=4.37) is consistent with prior research suggesting elevated PTSD risk in autism but provides more precise estimates from a population-based sample with prospective follow-up. ^4–6^ The association persisted after adjustment for parental psychiatric history, suggesting it is not fully explained by familial confounding, though residual confounding by unmeasured genetic or environmental factors cannot be excluded.

Several mechanisms may contribute to this heightened vulnerability. First, individuals with autism and ADHD may experience increased stress reactivity and emotional dysregulation. Research indicates that autistic individuals show stronger biological responses to stressors, amplified emotional responses, and poor emotional regulation compared to neurotypical individuals, with similar difficulties documented in ADHD populations.^16^ Increased stress reactivity and emotional dysregulation are established risk factors for PTSD and could account for vulnerability independent of trauma exposure. Second, individuals with neurodevelopmental conditions face chronic stress from multiple sources. Both autism and ADHD involve persistent difficulties with executive functioning that affect daily adaptive functioning. Such chronic stressor exposure is itself a risk factor for PTSD, potentially depleting psychological reserves needed to cope with acute traumatic events.^17^ Third, deficits in social cognition characteristic of autism, including inability to identify one’s own discomfort at inappropriate behavior and difficulty detecting violations of social norms, may increase vulnerability to interpersonal trauma and impede recovery through standard trauma processing mechanisms.^14^ When ADHD co-occurs with autism, impulsivity and inattention may further compromise the ability to recognize and respond appropriately to dangerous situations, compounding trauma risk. Whether these mechanisms operate with differing magnitude across sexes, and to what degree they account for the observed sex disparity, remains an important question for future research.

The identification of ADHD as an independent risk factor within autism has direct clinical implications. ADHD is common in autism (42.5% in our cohort) and is independently associated with PTSD risk.^11^ The combination produced particularly elevated risk: autistic females with ADHD had 6.0% 10-year cumulative incidence, corresponding to a NNS of only 18. This suggests that systematic PTSD screening in this subgroup could be efficient and clinically valuable. The elevated risk associated with ADHD may reflect shared vulnerability factors, increased trauma exposure related to impulsivity, or reduced capacity to employ coping strategies.

Perhaps most clinically significant are our findings regarding post-PTSD course. Autistic individuals with PTSD had more healthcare visits, higher hospitalization rates, and more persistent care needs than controls with PTSD. Nearly one quarter of autistic individuals with PTSD had ongoing care contacts in all three years following diagnosis, compared with only 12% of controls. This suggests that PTSD in autism may be more treatment-resistant, more severe, or both. Standard PTSD treatments may be less effective for autistic individuals, who may face barriers including communication differences, sensory sensitivities in treatment settings, and limited availability of autism-adapted interventions.^35,36^ The lower PTSD incidence among autistic individuals with intellectual disability warrants careful interpretation. While this could reflect genuine protective effects, it more likely reflects diagnostic challenges. Individuals with intellectual disability may have difficulty articulating PTSD symptoms, leading to diagnostic overshadowing where symptoms are attributed to the intellectual disability rather than recognized as PTSD.^37^ These findings support the need for autism-specific PTSD treatment approaches and sustained clinical follow-up beyond what might be typical for non-autistic patients.

Several limitations should be considered. First, PTSD ascertainment relied on clinical diagnoses recorded in specialist care, potentially missing cases treated exclusively in primary care or those who did not seek treatment. This would underestimate absolute incidence but is unlikely to bias the autism-control comparison unless healthcare-seeking differed systematically between groups. Additionally, our reliance on ICD-10 diagnostic codes may have missed cases where PTSD symptoms manifested atypically in autistic individuals, such as through developmental regression or increased autism-specific behaviors rather than classic PTSD symptomatology.^38,39^ Second, we lacked information on trauma exposure, preventing analysis of whether elevated risk reflects increased exposure, heightened vulnerability to similar exposures, or both. However, evidence suggests both mechanisms are operative: autistic individuals experience higher rates of trauma exposure and show heightened vulnerability to PTSD conditional on exposure.^14,34^ Third, the study period predated widespread recognition of autism-PTSD comorbidity, potentially affecting diagnostic practices. Fourth, Swedish registers do not capture PTSD symptom severity, precluding dimensional analysis. Fifth, although we adjusted for parental psychiatric history, residual confounding by shared genetic liability or unmeasured environmental factors cannot be excluded.

## CONCLUSION

In this population-based matched cohort study, autism was associated with substantially elevated risk of incident PTSD, particularly among females and those with comorbid ADHD. Ten-year cumulative incidence reached 6.0% among autistic females with ADHD, representing approximately 1 in 17 individuals. When PTSD occurred, autistic individuals experienced more severe and persistent clinical courses, with higher rates of psychiatric hospitalization and sustained care needs. These findings support routine PTSD screening in autism care, with prioritization of females and those with ADHD comorbidity. Autistic individuals who develop PTSD may require more intensive and sustained treatment approaches than typically provided to non-autistic patients, with particular attention to atypical symptom presentation including developmental regression. Further research is needed to develop and validate autism-adapted trauma assessment tools and treatment interventions.

## Data sharing statement

Data used in this study are from Swedish national registers and are not publicly available due to Swedish data protection regulations. Requests for data access should be directed to the Swedish National Board of Health and Welfare and Statistics Sweden.

## Financial Disclosure

None.

## Author Contributions

- Study concept and design: VB, BM, SS
- Acquisition, analysis, or interpretation of data: VB, BM, SS
- Drafting of the manuscript: All authors
- Critical revision of the manuscript for important intellectual content: All authors
- Statistical analysis: BM, SS
- Obtained funding: BM
- Study supervision: BM

## Funding/Support

This study was supported by a grant from the Beatrice and Samuel A. Seaver Foundation (BM, SS).

## Data Availability

Data may be obtained from a third party and are not publicly available. Data cannot be shared
publicly owing to restrictions by law. Data are available from the National Medical Registries in
Sweden after approval by the Swedish Ethical Review Authority.

## Notes

### Competing Interest Statement

The authors have declared no competing interest.

### Author Declarations

The Regional Ethical Review Board in Stockholm approved this study (Dnr 2013/862-31/5). Swedish law permits register-based research without individual consent when data are anonymized.

