## Supplementary Tables for "Sex Differences in PTSD Risk Among Autistic Individuals: A Population-Based Matched Cohort Study"

**Supplemental Table 1.** Exploratory Analyses: Age at Diagnosis and Autism Subtype

**1A.** Age at Autism Diagnosis and PTSD Risk

| Analysis | No. | Events | HR (95% CI) | P |
|---|---|---|---|---|
| **Age at diagnosis (categorical)** | | | | |
| 6-9 years (ref) | — | — | 1 [Reference] | — |
| 10-12 years | — | — | 1.12 (0.67-1.89) | .66 |
| 13-15 years | — | — | 1.93 (1.21-3.09) | .006 |
| 16+ years | — | — | 2.35 (1.48-3.72) | <.001 |
| **Age at diagnosis, per year** | | | | .87 |
| Overall | 42,866 | 401 | 1.09 (1.06-1.13) | <.001 |
| Males | 28,851 | 93 | 1.09 (1.03-1.16) | .005 |
| Females | 14,015 | 308 | 1.09 (1.05-1.13) | <.001 |
| **Sex difference attenuation** | | | | |
| Female HR, unadjusted | — | — | 6.63 (5.26-8.37) | <.001 |
| Female HR, adjusted for age at diagnosis | — | — | 6.15 (4.87-7.77) | <.001 |
| Percent attenuation | — | — | 4% | — |

**1B.** Asperger Syndrome vs Other Autism and PTSD Risk

| Analysis | No. Asperger | No. Other | Events Asperger | Events Other | HR (95% CI) | P |
|---|---|---|---|---|---|---|
| Adjusted for sex and birth year | 17,136 | 25,730 | 219 | 182 | 1.33 (1.09-1.62) | .005 |
| Fully adjusted | 17,136 | 25,730 | 219 | 182 | 1.16 (0.95-1.42) | .15 |
| **Sex-stratified (fully adjusted)** | | | | | | .45 |
| Males | 11,245 | 17,606 | 46 | 47 | 1.17 (0.77-1.76) | .47 |
| Females | 5,891 | 8,124 | 173 | 135 | 1.38 (1.10-1.73) | .005 |

Abbreviations: HR, hazard ratio. 1A analyses restricted to autistic individuals; P value in "per year" header row is for sex × age interaction. 1B reference group is other autism diagnoses (ICD-10 F84.0, F84.1, F84.8, F84.9); P value in "sex-stratified" header row is for sex × subtype interaction. All models stratified by matched set with robust standard errors clustered on the biological mother.

**Figure S1.** Cumulative Incidence of PTSD Among Autistic Individuals and Matched Controls. Cumulative incidence curves showing the proportion of individuals diagnosed with posttraumatic stress disorder (PTSD) over time since the index date. (A) Overall comparison between autistic individuals and matched controls. (B) Stratified by sex. (C) Stratified by ADHD status. Shaded areas represent 95% confidence intervals. P values are from the log-rank test.

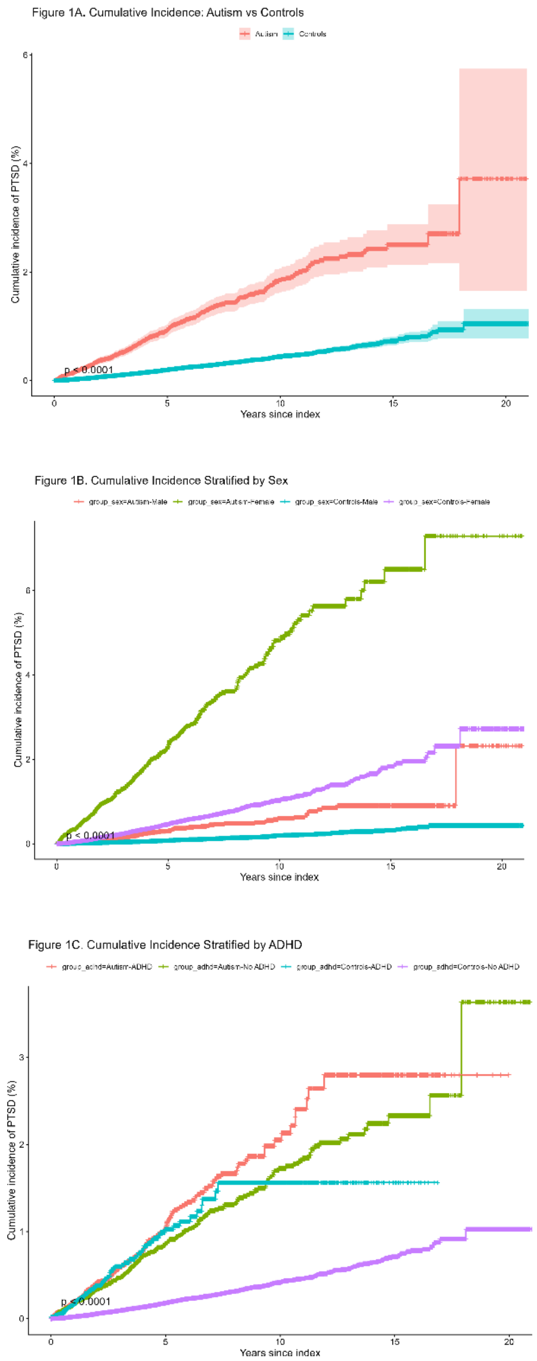


**Figure S2.** Cumulative incidence curves for posttraumatic stress disorder (PTSD) stratified by sex and attention-deficit/hyperactivity disorder (ADHD) comorbidity among autistic individuals. Shaded areas represent 95% confidence intervals.

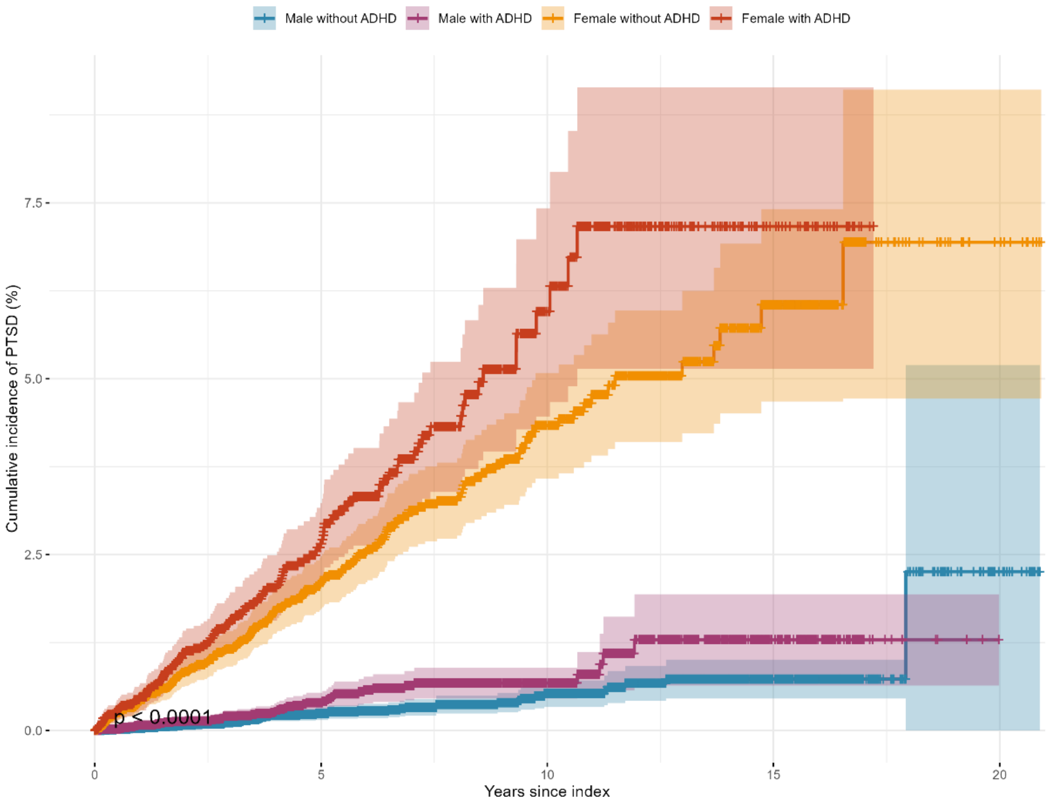

Male without ADHD
Male with ADHD
Female without ADHD
Female with ADHD
Cumulative incidence of PTSD (%)
0.0
2.5
5.0
7.5
p < 0.0001
0
5
10
15
20
Years since index
